# Longitudinal Clinical Foundation Models Augmented with Genomics for Early Detection and Risk Stratification of Inherited Cardiomyopathy

**DOI:** 10.64898/2026.08.10.26360107

**Authors:** Andrew Zolensky, Colleen M. Kripke, Karl Keat, Scott M. Damrauer, Michael G. Levin, Anurag Verma

## Abstract

Hypertrophic and dilated cardiomyopathy (HCM and DCM) carry substantial morbidity and mortality, yet diagnosis may be delayed, particularly when presentation is nonspecific. Existing machine-learning approaches to cardiomyopathy phenotyping, genotype prediction, and risk stratification commonly rely on disease-specific, hand-engineered features drawn from echocardiography, cardiac MRI, ECG, or curated clinical variables. We evaluated whether a general-purpose clinical foundation model, CLMBR-T-base, pre-trained via next-clinical-event prediction with no cardiomyopathy-specific supervision, could produce linearly separable embeddings for all three case/control cohorts. Using EHR data from the Penn Medicine BioBank, we constructed cohorts for (1) prediction of a first recorded qualifying HCM/DCM diagnosis at 1-, 3-, and 6-month horizons, decomposed into eventual-versus-never-case and imminent-versus-eventual comparisons; (2) genetic carrier status prediction among diagnosed patients with completed gene panels; and (3) prediction of heart-failure hospitalization, and all-cause mortality as both binary and time-to-event outcomes. Linear probes fitted to frozen embeddings achieved AUROCs of 0.75-0.82 for onset prediction, 0.74-0.75 for genotype status, and Harrell’s concordance of 0.65-0.80 for time-to-event outcomes. Decomposing the onset prediction task reveals that the model often misclassifies patients who were diagnosed later as positive, suggesting the patient journey embeddings encode disease state more reliably than care timing. These results suggest that a single, generically pretrained EHR embedding can support multiple clinically motivated prediction problems in CM without disease-specific feature engineering.

## 1. Introduction

Cardiomyopathies are disorders of the myocardium that can lead to heart failure, arrhythmia, thromboembolism, and sudden cardiac death.^1,2^ HCM is commonly estimated to affect approximately 1 in 500 people.^1^ DCM prevalence estimates vary with case definition and ascertainment. HCM is characterized by otherwise-unexplained left-ventricular hypertrophy and may be obstructive or nonobstructive. DCM is characterized by left-ventricular or biventricular dilation and systolic dysfunction not explained solely by abnormal loading conditions or coronary disease.^2^ Both disorders can have genetic causes, although DCM is etiologically heterogeneous and many patients do not receive a molecular diagnosis.^3^

While HCM and DCM patients face substantial morbidity, multiple treatment options exist, including exercise therapy, medications, and surgical interventions,^4,5^ and early detection can allow time for risk stratification to be performed and targeted intervention to be made before irreversible remodeling occurs.^6,7^ But despite the value of early intervention, HCM and DCM are often mistaken by clinical professionals for other cardiovascular disorders.^8^ In particular, studies estimate that more than half of patients with obstructive HCM receive a delayed diagnosis, and that patients have an average of 4 misdiagnoses prior to the correct diagnosis being made.^8^ Delays in diagnosis can be largely attributed to many patients being asymptomatic in addition to costs associated with the testing procedures required to establish the disease, which include cardiac imaging to assess maximum LV wall thickness and in many cases genetic testing to assess carrier status.^8^

A substantial statistics and machine-learning literature has therefore emerged to address the three problems of early detection, carrier identification, and risk stratification in isolation, almost always using disease-specific, hand-engineered feature sets drawn from echocardiography, cardiac MRI, ECG waveforms, or curated clinical variables (Section 2). Comparatively, little work has asked whether a single general-purpose representation of a patient’s longitudinal structured EHR, learned once, without any cardiomyopathy-specific supervision, and reused across many downstream tasks, can support all three judgments at once. Despite this, performant tools fitting this description could decrease production time for predictive health analytics systems, or alternatively provide a generic prior in a mixture of experts model which takes specific modality information into account. Clinical foundation models trained via next-event prediction on large volumes of coded EHR data are designed precisely for this kind of transfer learning: a small, cheap-to-train prediction head is fit on top of frozen patient-trajectory embeddings for a specific downstream task. This paradigm has shown promise for general administrative and health-state-forecasting benchmarks, but it has not, to our knowledge, been evaluated on a set of tasks defined by domain experts around a specific rare genetic disease and linked to genomic ground truth.

CLMBR-T-Base^9^ is an open-source, autoregressive transformer based on the CLMBR architecture^10^ and trained via next-clinical-event prediction over sequences of OMOP-standardized medical codes. It was pretrained on 2.57 million de-identified Stanford Medicine patients as part of the EHRSHOT benchmark.^9^ The model can be used along with the associated FEMR library to convert longitudinal patient records into fixed-length patient embeddings, which can then serve as input to downstream linear classifiers or Cox models; a transfer-learning approach known as linear probing.^11^ EHRSHOT’s own evaluation tasks are intentionally general and are not designed to test rare, genetically defined cardiomyopathies. In this study, we used CLMBR-T-Base, operating on EHR data from the Penn Medicine BioBank (PMBB) harmonized to the OMOP Common Data Model and converted to the Medical Event Data Standard (MEDS),^12^ to construct and evaluate three families of prediction tasks for HCM and DCM. Our contributions are as follows:

1. We construct task-definition logic for three clinically motivated HCM/DCM prediction problems built directly on OMOP structured data and converted into the MEDS format, designed to be reproducible across OMOP-mapped sites.
2. We explicitly decompose the onset-prediction task into an “eventual case versus never-case” comparison and a harder “imminent versus delayed presentation among eventual cases” comparison, revealing that most of the apparent discriminative power of the model reflects the former rather than the latter.
3. We report our results on HCM/DCM early detection, gene carrier status prediction, adverse event prediction, and time to adverse event prediction, quantitatively and where possible with matched confidence intervals, and compare against the published literature on HCM/DCM diagnosis, genotype, and outcome prediction.

## 2. Related Work

### 2.1. Structured-record-based diagnosis and onset prediction

Automated identification of HCM/DCM cases from structured EHR data has mostly targeted case ascertainment rather than prediction of the time to the first recorded qualifying diagnosis from structured EHR data. Farahani et al. trained a random-forest classifier on ICD-10 billing-code histories from the Mayo Clinic EHR to distinguish “definite,” “possible,” and “no” HCM.^13^ Other studies have attempted early detection of HCM and DCM by focusing on specific modalities, such as digitized ECG signals or images.^14^ These studies primarily addressed case ascertainment or used disease-specific modalities rather than reusable longitudinal EHR representations..

### 2.2. Genetic carrier status prediction

Previous studies have evaluated automated prediction of genotype-positive status for HCM and DCM patients. For HCM, Liang et al. trained machine-learning models on readily available clinical and echocardiographic variables to predict sarcomeric genotype positivity, achieving an AUROC of 0.92.^15^ Captur et al. showed that a small set of cardiac MRI structural features discriminates sarcomere-mutation carriers among subclinical, phenotype-negative relatives (AUC 0.85).^16^ Zhou et al. trained a deep-learning model on cardiac cine-MRI images to classify HCM genotype, and Morita et al. combined a deep convolutional neural network over echocardiographic images with the Mayo HCM Genotype Predictor score (AUC 0.86).^17–19^ For DCM, Escobar-López et al. derived and externally validated the “Madrid Genotype Score,” a five-variable clinical/ECG model that predicts pathogenic-genotype positivity with a C-statistic of 0.75.^20^ All of these approaches rely on manually curated clinical, ECG, or imaging variables selected by domain experts; we did not identify a study evaluating a general-purpose EHR foundation-model representation for this task.

### 2.3. Time-to-outcome and survival prediction

Geyer et al. fit a Cox model with elastic-net regularization on 33 clinical, genetic, echocardiographic, and CMR variables accross 604 HCM patients, achieving Harrell’s concordance of 0.75 for major adverse cardiac events (MACEs).^21^ Zhao et al. trained a LightGBM model on 14 CMR imaging variables and 23 clinical features in a four-center cohort of 758 HCM patients, achieving AUC 0.83 for SCD prediction.^22^ Lai et al. trained a multimodal transformer, MAARS, on EHR text, radiology reports, and CMR images in HCM patients, achieving AUC 0.89/0.81 for arrhythmic sudden death.^23^ For DCM, Shu et al. reported AUC 0.87 for adverse-event prediction using CMR imaging texture features, and MacGregor et al. achieved AUC 0.94 for a DCM therapy non-responder prediction task based on MRI imaging features.^24,25^ Corian`o et al. trained a deep-learning survival model (DARP-D) on 154 DCM patients’ cine/LGE CMR plus clinical covariates, and report a 95% CI for Harrell’s C of 0.12–0.68, illustrating that rich imaging input does not guarantee good performance in small-cohort DCM survival modeling.^26^ Collectively, these models use disease-specific imaging, ECG, or curated clinical features and generally evaluate arrhythmic or composite outcomes. Differences in cohorts and endpoint definitions limit direct comparison with the present study.

## 3. Methods

### 3.1. Study Design and Data Source

We conducted a retrospective, single-site prediction-model development and internal evaluation study using structured electronic health record and whole-exome sequencing data from the Penn Medicine BioBank (PMBB).^27^ PMBB is an EHR-linked, OMOP-v5.3-compliant database with 365,269 patients as of April 2026. Of these patients, 70,925 have had whole exome sequencing for research use, with coverage of cardiomyopathy-associated genes listed in Table 1 below. Patient event were extracted from OMOP visit, measurements, observations, procedures, conditions, medications, person, and death, in order to extract patient events. All task families were constructed and modeled separately for HCM and DCM using an identical overall pipeline.

**Table 1.** Genes used to define inclusion and exclusion control for HCM and DCM cases and controls.

| Evidence tier | HCM genes | DCM genes |
| --- | --- | --- |
| Positive | MYBPC3, MYH7, TNNI3, TNNT2, TPM1, MYL2, MYL3 | TTN, LMNA, MYH7, TNNT2, BAG3, RBM20, FLNC, DSP, PLN |
| Limited/Uncertain | ALPK3, CSRP3, FLNC, TRIM63, PRKAG2 | DES, CSRP3, TRIM63, SCN5A |
| Negative | DES, DSP, PLN, LMNA, BAG3, RBM20, SCN5A, TTN, FBN1 | MYBPC3, TNNI3, TPM1, MYL2, MYL3, ALPK3, PRKAG2, FBN1 |

### 3.2. Data representation and embedding extraction

Structured OMOP-CDM data were converted to the MEDS (v0.1.3) format using the medsetl toolchain (v0.1.3), and processed into patient timelines using FEMR (v0.2.3), from which CLMBR-T-base patient-trajectory embeddings were generated. Eligible patients were required to have at least two visits in PMBB recorded after their EHR start date.

Formally, each task is represented using the MEDS LabelSchema.^12^ Each example contains a patient id, a prediction (cutoff) time, and a label. Correspondingly, each patient is represented using the MEDS DataSchema,^12^ in which a patient is represented as a sequence of events, each referencing a patient id, a concept code (OMOP concept id), a timestamp, and a text or numeric value if applicable. Events include measurements, diagnoses, medication orders, procedures, and other observations. Special “events” are assigned for an individual’s birth and demographics, with the timestamp set to the birth date in order to bring these items to the beginning of the sequence.

### 3.3. Prediction task and cohort creation

We evaluated three task families separately for HCM and DCM: onset prediction of the first recorded qualifying diagnosis, genotype-status prediction, and prediction of post-diagnosis clinical outcomes. Our overall approach is depicted in Figure 1.

**Fig. 1.**
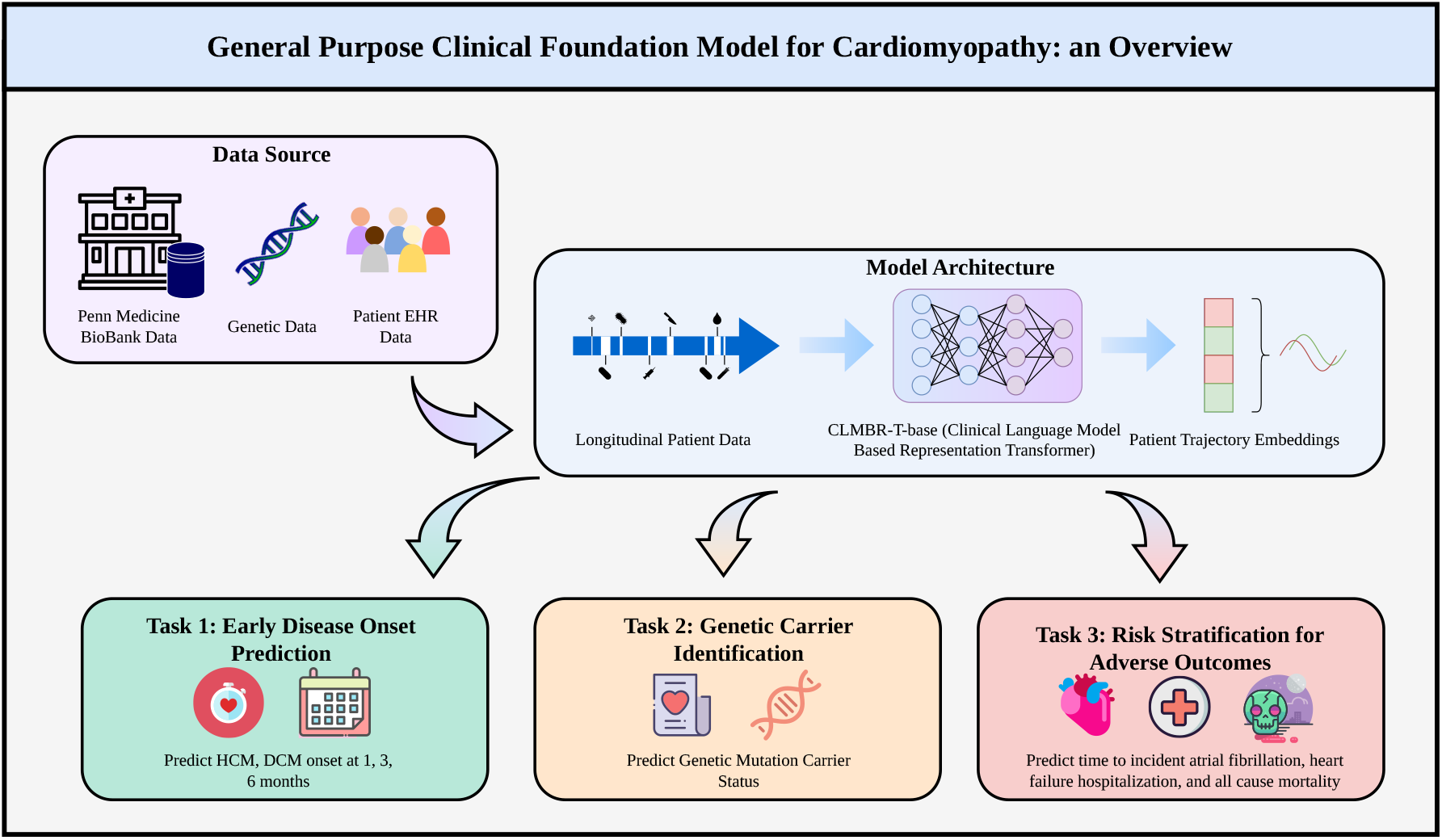
Process Diagram of the methodology employed in this paper.

We use CLMBR-T-base embeddings of the patient trajectory up to the prediction time to predict the label in question. Our experiments are as follows:

1. Predict whether an HCM/DCM diagnosis is imminent (1 month, 3 months, 6 months), anchoring on a date prior to diagnosis
2. Predict whether an HCM/DCM patient is a genetic carrier or not, anchoring on the first diagnosis date
3. Predict outcomes associated with HCM/DCM and the time to event, anchoring on the first diagnosis date

For our third experiment,heart failure hospitalization (HFH) was modeled as a binary and time-to-event outcome, while all-cause mortality was modeled as a time-to-event outcome.

#### 3.3.1. Disease Onset Task

The disease onset task predicts if the patient will receive their first confirmed HCM or DCM diagnosis within a pre-specified time interval or not. Three time windows were used to define task cohorts: 1 month, 3 months, and 6 months, each tested separately for HCM and DCM. A confirmed diagnosis was defined as a qualifying pair: a transthoracic echocardiogram (CPT-4 93306, 93307, or 93308) occurring within 90 days of a diagnosis phecode for HCM (CV 414.1, CV 414.11) or DCM (CV 414.2). The diagnosis date for the patient was the earliest date in any qualifying pair. For cases, the prediction cutoff date was the end time of a random visit concluding no more than the specified number of months before the first diagnosis date. For patients with a qualifying pair, the prediction cutoff date was the end time of a random visit ending more than the specified amount of time before the first diagnosis. For patients without a qualifying pair, the prediction cutoff date was the end time for a random visit. The control group therefore contained a group of patients who never developed the condition in question, and a group of patients who developed it too late to fall within the specified time interval. For purposes of analysis, the cases are referred to as “immanent converters,” whereas the subset of controls who eventually receive the diagnoses are referred to as “eventual converters.”

#### 3.3.2. Genotype Status Task

Cardiomyopathy-associated genes were prespecified based on ClinGen gene–disease validity classifications (citation) and are listed in Table 1. Genotype-positive status was defined as carrying at least one qualifying finding in a Positive-tier gene for the corresponding cardiomyopathy. Patients with a completed panel showing no qualifying finding in a Positive-tier or Limited/Uncertain-tier gene were labeled genotype-negative. Patients with a finding in a Limited/Uncertain-tier gene, or an incompletely assessed panel, were excluded.

#### 3.3.3. Post-diagonsis outcomes task

Using the first qualifying HCM or DCM diagnosis as the prediction cutoff time, we defined two progression outcomes: Heart-Failure Hospitalization (HFH), and all-cause mortality. HFH was modeled as both a binary classification and time-to-event task, whereas mortality was modeled as a time-to-event task.

For time-to-event analyses, follow-up began at the qualifying date and continued until the first occurrence of the outcome or the last available visit date on record for that patient. HFH was defined by matching visits with a hospitalization to phecodes for heart failure, and all-cause mortality was defined using the OMOP death table.

### 3.4. Models and preprocessing

We used the pre-trained CLMBR-T-base model to embed our patient histories before fitting linear regression models using the embedding dimensions (or their dimensionality-reduced versions) as covariates, an approach often referred to as “linear probing.” We preserve only the final hidden state embedding of the most recently seen event for a given patient/cut-off date combination, as by the final layer this has been updated through several layers of attention to encode the information about the sequence of events up to this point that is needed to predict the next event. Additionally, to reduce the amount of compute burden, we truncate patient histories prior to embedding to only the most recent 100 coded events, leaving room for demographics events at the start of the sequence. This yields a single 768-dimensional vector as our covariates representing a given patient trajectory up to a cutoff time (the prediction cutoff date).

For binary event prediction, we utilize logistic regression to predict the associated event probability. We utilize L2 regularization during fitting, and tune the regularization strength, *C*, via grid-search over the training set. For genotype prediction in particular, we also utilize PCA with a number of principal components determined via grid search over the training set, due to the small sample size. For survival analysis, we use Cox proportional hazards models to estimate the hazard function, and fit our models to the leading 20 principal components of the embeddings.

### 3.5. Model Evaluation

For all binary classification experiments we report AUROC (95% CI), PR-AUC (95% CI), and 0.5-thresholded sensitivity and specificity on a held-out test subset consisting of 20% of the original patient cohort (80/20 train/test split on patient id). For the time to event experiments we report Harrell’s Concordance (95% CI).

### 3.6. Compute, reproducibility

All computation was performed on a single-node Databricks cluster (Standard D32ds v5, 32 vCPU, Spark 17.3.x) provisioned within the Penn Medicine BioBank secure compute environment. Data engineering and cohort construction were implemented in PySpark using a custom Python package, meds-biobank (available on PyPI: https://pypi.org/project/meds-biobank/), which reimplements the MEDS-ETL (v0.1.3) transformation logic for execution on Databricks against OMOP-formatted BioBank data. Patient timelines conforming to the Medical Event Data Standard (MEDS v0.1.3) were subsequently processed using FEMR (v0.2.3) to generate CLMBR-T-base patient-trajectory embeddings. All task-cohort construction, modeling, and evaluation code is available at https://github.com/AndrewLZolensky/Multi-Omic-Augmentation.

## 4. Results

### 4.1. Cohort characteristics

The cohort characteristics are broken down in 2. Columns are provided for the cohort as a whole and just the patients with whole-exome sequencing performed. At the bottom, the size of the HCM, DCM, HCM gene positive, and DCM gene positive cohorts is shown.

| Characteristics | PMBB |  |
| --- | --- | --- |
|  | All Consented | Genotyped/WES |
| **Cohort size** |  |  |
| Participants, n | 365,269 | 70,925 |
| **Demographic characteristics** |  |  |
| EHR-reported race and ethnicity, n (%) |  |  |
| White, n (%) | 259,152 (70.9) | 49,518 (70.6) |
| Black, n (%) | 54,806 (15.0) | 14,053 (20.0) |
| Asian, n (%) | 16,238 (4.4) | 2,023 (2.9) |
| Hispanic, n (%) | 15,221 (4.2) | 2,082 (3.0) |
| Female sex, n (%) | 204,858 (56.1) | 36,237 (51.7) |
| **Cardiovascular phenotype** |  |  |
| Hypertrophic Cardiomyopathy (I42.1, I42.2) | 1,736 (0.005) | 665 (0.009) |
| Dilated Cardiomyopathy (I42.0, I43) | 2,895 (0.008) | 1,441 (0.020) |
| HCM Gene Positive | - | 352 (0.005) |
| DCM Gene Positive | - | 528 (0.007) |

### 4.2. Disease onset prediction

Table 3 and Table 4 present the results for Experiment 1. Table 3 breaks down the results by disease category (HCM/DCM), then lead time (1mo, 3mo, 6mo). The number of patients in the training and testing categories, as well as the balance of cases to controls, is also shown. For each combination of a disease and lead-time, the AUROC (95% CI), PR-AUC (95% CI), and 0.5-thresholded sensitivity and specificity are provided. AUROC remains consistently above PR-AUC in all experiments, while sensitivity and specificity remain balanced. No clear trend is observed when considering increasing lead times. Table 4 shows the results for Experiment 1 restricted to include only imminent and eventual converters. While AUROC declined from the full setting reported in Table 3, PR-AUC is seen to increase relative to that condition. Additionally, while sensitivity remains similar, specificity decreases substantially relative to the full design.

**Table 3.** Results for Experiment 1: Onset Prediction at lead times of 1, 3, and 6 months, for HCM and DCM.

| Dis. | Lead | n (train) | n (test) | AUROC (CI) | PR-AUC (CI) | Sen./Spec. |
| --- | --- | --- | --- | --- | --- | --- |
| HCM | 1 mo | 4513 (1139/3374) | 1141 (291/850) | 0.758 (0.730, 0.786) | 0.495 (0.451, 0.549) | 0.68/0.70 |
| HCM | 3 mo | 4726 (1202/3524) | 1174 (293/881) | 0.765 (0.739, 0.792) | 0.480 (0.437, 0.534) | 0.71/0.70 |
| HCM | 6 mo | 4804 (1192/3612) | 1222 (316/906) | 0.752 (0.724, 0.778) | 0.502 (0.455, 0.549) | 0.67/0.69 |
| DCM | 1 mo | 4969 (1239/3730) | 1240 (322/918) | 0.802 (0.776, 0.823) | 0.564 (0.520, 0.610) | 0.71/0.73 |
| DCM | 3 mo | 4920 (1192/3728) | 1248 (306/942) | 0.822 (0.799, 0.843) | 0.593 (0.547, 0.642) | 0.73/0.73 |
| DCM | 6 mo | 4961 (1275/3686) | 1221 (308/913) | 0.804 (0.782, 0.826) | 0.535 (0.494, 0.584) | 0.68/0.76 |

**Table 4.** Results for Experiment 1: Onset Prediction (converters only, imminent vs. eventual converters), at lead times of 1, 3, and 6 months, for HCM and DCM. Models are the same as in the full-task table, re-scored on the converter subset.

| Disease | Lead | n (test), pos/neg | AUROC (95% CI) | PR-AUC (95% CI) | Sens./Spec. |
| --- | --- | --- | --- | --- | --- |
| HCM | 1 mo | 582 (291/291) | 0.673 (0.6272, 0.7162) | 0.657 (0.5961, 0.7215) | 0.68/0.56 |
| HCM | 3 mo | 582 (289/293) | 0.662 (0.6173, 0.7073) | 0.643 (0.5791, 0.7045) | 0.71/0.56 |
| HCM | 6 mo | 626 (310/316) | 0.660 (0.6167, 0.6997) | 0.658 (0.6023, 0.7114) | 0.67/0.53 |
| DCM | 1 mo | 645 (323/322) | 0.655 (0.613, 0.697) | 0.642 (0.5853, 0.7054) | 0.71/0.49 |
| DCM | 3 mo | 651 (345/306) | 0.678 (0.637, 0.717) | 0.651 (0.595, 0.708) | 0.73/0.49 |
| DCM | 6 mo | 603 (295/308) | 0.643 (0.6009, 0.6850) | 0.618 (0.5600, 0.6780) | 0.68/0.53 |

Table 5 presents the results for Experiment 2. Compared to Table 3, Table 5 has an additional column for the number of principal components retained in a dimensionality-reduction sensitivity analysis. In particular, for both HCM and DCM sub-experiments, we repeat our design using PCA, with the number of principal components selected by grid search. The resulting number of PC’s is given in the column, “nPCs.” Table 5 shows an imbalance between AUROC and PR-AUC, with the AUROC’s for HCM and DCM as 0.745 and 0.744 respectively, and their PR-AUC’s trailing behind at 0.478 and 0.601, respectively. Specificity was higher than sensitivity for both HCM and DCM. Grid search for HCM identifies 100 principal components as optimal, the upper limit of the evaluated range. DCM identifies 30 as optimal, and the PCA-reduced model had lower observed test performance than the unreduced model.

**Table 5.**
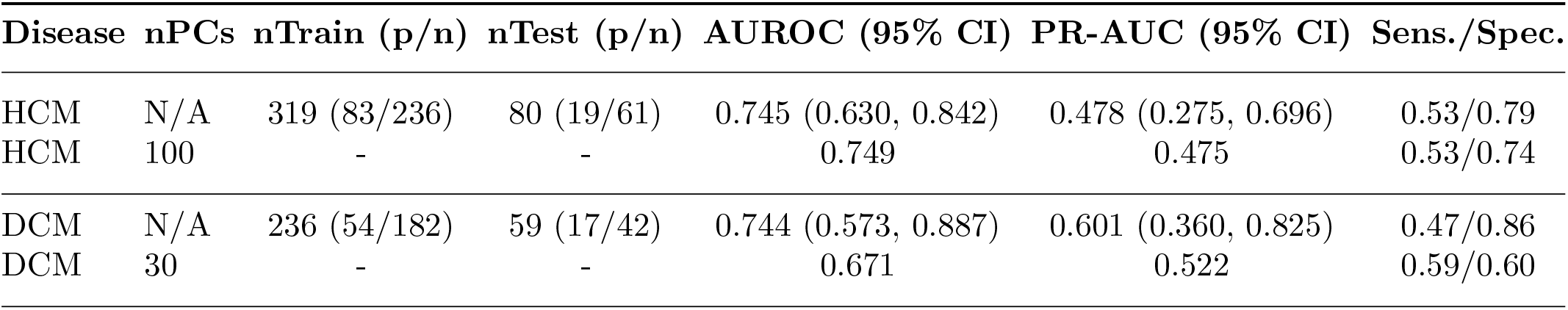
Results for Experiment 2: gene carrier status prediction. Includes results for feature ablation: CLMBR embeddings with and without PCA dimensionality reduction, for HCM and DCM.

### 4.3. Post-diagnosis outcomes

Table 6 and Table 7 present the results for Experiment 3. Table 6 covers binary outcome prediction and Table 7 covers time-to-event prediction. Fior binary outcomes, the results for DCM are balanced across AUROC and PR-AUC, with a slightly higher PR-AUC, whereas the trend for HCM is reversed and slightly more pronounced. Sensitivity and specificity also remain balanced, although specificiy tends to be higher.

**Table 6.** Results for heart failure hospitalization, for HCM and DCM.

| Disease | n (train) | n (test) | AUROC (95% CI) | PR-AUC (95% CI) | Sens./Spec. |
| --- | --- | --- | --- | --- | --- |
| DCM | 1350/966 | 331/248 | 0.690 (0.646, 0.735) | 0.719 (0.6690, 0.7714) | 0.66/0.66 |
| HCM | 475/913 | 134/214 | 0.678 (0.626, 0.732) | 0.535 (0.460, 0.621) | 0.62/0.65 |

**Table 7.**
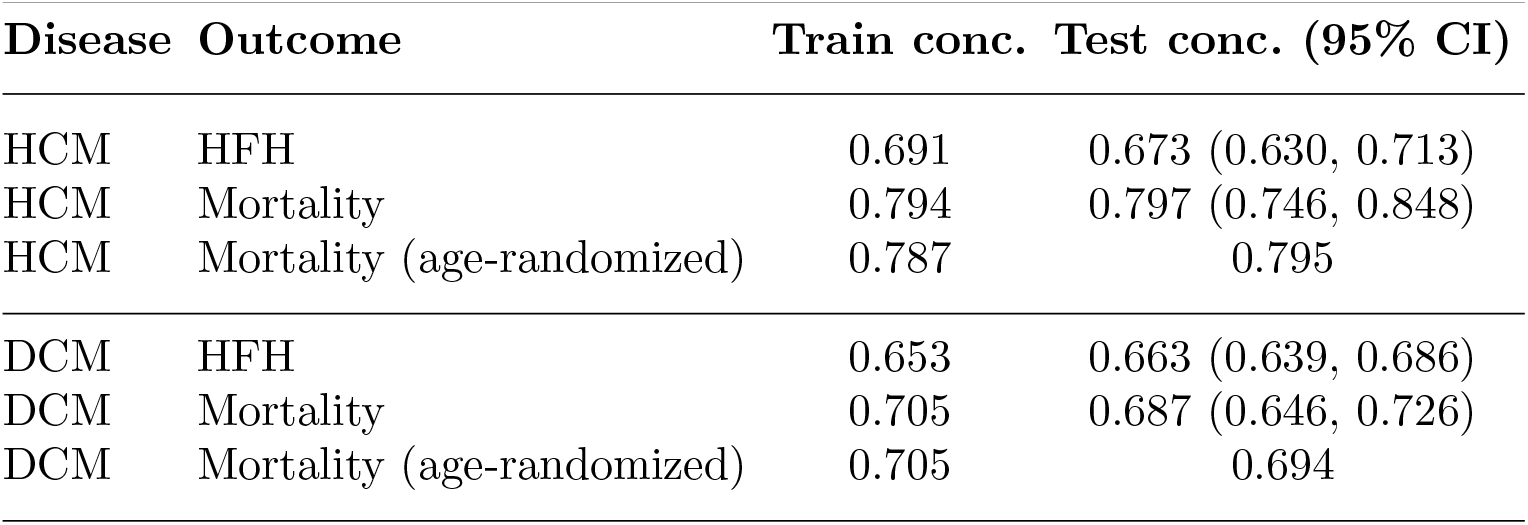
Cox proportional-hazards concordance for time-to-progression out-comes.

| Disease | Outcome | Train conc. | Test conc. (95% CI) |
| --- | --- | --- | --- |
| HCM | HFH | 0.691 | 0.673 (0.630, 0.713) |
| HCM | Mortality | 0.794 | 0.797 (0.746, 0.848) |
| HCM | Mortality (age-randomized) | 0.787 | 0.795 |
| DCM | HFH | 0.653 | 0.663 (0.639, 0.686) |
| DCM | Mortality | 0.705 | 0.687 (0.646, 0.726) |
| DCM | Mortality (age-randomized) | 0.705 | 0.694 |

The adverse events for which performance is reported in Table 7 consist of Heart Failure Hospitalization (HFH) and all-cause mortality. The table reports Harrell’s Concordance (C) on the training set and the testing set (95% CI). Training and test concordance estimates were generally similar, although test concordance was lower for some outcomes for HCM-HFH, and DCM-Mortality. Randomizing the explicit age input produced little change in test concordance.

## 5. Discussion

This study evaluated whether frozen representations from a general-purpose clinical next-event model can be used to predict onset, genotype status, outcomes form DCM and HCM. First, we observed that discrimination was stronger for distinguishing eventual cases from patients without a recorded qualifying diagnosis than for distinguishing imminent from later diagnosis among eventual cases. Second, genotype-status discrimination was significant, although sensitivity was limited at the evaluated threshold in the low-regime data. Third, the embeddings contained prognostic information for post-diagnosis outcomes, with the strongest observed concordance for mortality among patients with HCM. These findings support further evaluation of reusable structured-EHR representations.

### 5.1. Onset prediction and the eventual/imminent decomposition

Table 3 reveals moderate AUROC scores (0.75–0.82) along with balanced sensitivity/specificity (0.67–0.76). This pattern indicates moderate discrimination, while the lower AUPRC (0.48–0.59) partly reflects outcome prevalence (1:3 p/n). The converters-only analysis (Table 4) clarifies where much of that discriminative power actually comes from. AUROC drops substantially (e.g., HCM 1mo: 0.758 *→* 0.673) while PR-AUC rises (0.495 *→* 0.657), consistent with the higher, 50% base rate in this subset. Critically, sensitivity is nearly unchanged while specificity collapses. This means “eventual converters” are more difficult negative examples for the model to separate from”imminent converters.” Much of the full-cohort AUROC therefore reflects separating eventual cases from patients without a recorded qualifying diagnosis during observed follow-up, rather than precisely distinguishing imminent from later recorded diagnosis. Also, this endpoint represents the first recorded qualifying diagnosis rather than biological disease onset. Differences in healthcare utilization, observation time, and diagnostic workup may therefore contribute to model performance.

### 5.2. Genotype status prediction

The AUROC score remains moderate here but PR-AUC is significantly lower than in Experiment 1, and sensitivity/specificity are markedly asymmetric rather than balanced as in Experiment 1. At a threshold of 0.5, the model had lower sensitivity than specificity for genotype-positive status. Relative to the literature, CLMBR-T-base embeddings trail disease-specific models built on curated echo/MRI features for HCM although direct numerical comparison is limited by differences in cohorts, predictors, and genotype definitions. For DCM the results are comparable to previous disease-specific models, despite using no hand-selected clinical or ECG variables at all. The PCA robustness check reflects the small sample size: HCM’s grid search saturates at 100 components (the upper bound tested), suggesting the search is overfitting to a training set of only 319 patients; DCM’s optimum at 30 components coincides with lower observed test performance (0.744/0.601 *→* 0.671/0.522), suggesting the reduced representation is discarding signal.

### 5.3. Outcome and survival prediction

Table 6 shows both moderate AUROC and PR-AUC for HFH when DCM patients are considered. For HCM patients, AUROC is moderate and PR-AUC is lower; this tendency is most pronounced for HFH. This is consistent with the outcomes differing in base rate across disease. In Table 6, Cox concordance is comparatively strong for HCM mortality (0.797). Re-running the HCM/DCM mortality models with patient age randomized prior to embedding produces no clear change in concordance, indicating the embeddings are not simply encoding chronological age as a proxy for mortality risk; rather, the prognostic signal appears to carry genuine information about the time to event. Modest train/test gaps for HCM-HFH, and DCM-Mortality indicate mild overfitting.

Across all three experiments, the trends in AUROC and PR-AUC track base positive rate: the gap between them widens where positives are rarest and narrows or reverses where positives are more common. Where it’s largest alongside decent sensitivity/specificity, it reflects a high absolute false-positive count per detected true positive. The consistent finding across tasks is that a single, disease-agnostic, next-event-pretrained embedding, fit only with a linear head, recovers a substantial fraction of the discriminative signal achieved by disease-specific, hand-engineered pipelines in the literature, without any HCM/DCM-specific feature construction.

## 6. Conclusion

We evaluated whether a general-purpose clinical foundation model embedding, pretrained via next-event prediction with no cardiomyopathy-specific supervision, could, via linear probing alone, support three distinct clinically-motivated problems in HCM and DCM: early detection, genetic carrier status prediction, and adverse-event/survival risk stratification. Across all three, the embeddings produced meaningful, statistically robust discrimination above chance. Performance trailed the best published disease-specific models built on curated echocardiographic, CMR, or ECG features, particularly for HCM genotype prediction, but was comparable to curated clinical-score approaches for DCM genotype prediction and produced non-trivial concordance for time-to-event outcomes, including mortality risk that survives an age-randomization control. Decomposing the onset-prediction task into eventual-vs-never-case and imminent-vs-eventual comparisons showed that most of the model’s apparent temporal foresight is in fact case ascertainment rather than fine-grained timing, a distinction with direct implications for how such models should be positioned in a clinical workflow (population-level screening versus urgent-referral triage). Taken together, these results support the feasibility of reusing a single, generically pretrained EHR embedding across multiple task families for a rare genetic disease area, reducing the need for redundant, disease-specific feature engineering at the cost of some performance relative to specialized models.

## 7. Limitations and Future Work

There are several limitations to this work. First, all cohorts were drawn from PMBB, a single academic health system’s biobank-linked EHR. Diagnosis rates, coding practices, and qualifying-pair definitions are institution-specific, while CLMBR-T-base was pretrained at Stanford Medicine. Performance may not transfer across demographics, referral patterns, or OMOP-mapping conventions; therefore, external, multi-site validation is needed. Second, the Experiment 2 genotype-prediction cohorts (319/80 and 236/59 train/test) were much smaller because completed genetic panels are rare. This limits confidence-interval precision, raises concerns about overfitting, and supports multi-site pooling or transfer approaches. Third, we evaluated only a frozen 768-dimensional final hidden-state embedding with a linear or PCA-reduced head. Fine-tuning, non-linear probes, and full-sequence pooling may improve performance and clarify whether limitations arise from the representation or the probe. Fourth, our case definition, echocardiography within 90 days of a diagnosis phecode, is a pragmatic proxy that may miss patients diagnosed through genetic testing, family screening, or outside records. Chart-review validation is therefore needed. Fifth, non-converter controls were sampled without matching on age, comorbidity, or healthcare utilization, while eventual converters represented a narrower, retrospectively sicker population. Matched-control or landmark-time analyses could separate model limitations from cohort-construction effects. Sixth, outcome definitions differed from prior literature, limiting direct comparison. We modeled heart-failure hospitalization, and all-cause mortality rather than arrhythmic or sudden-death composite outcomes. Finally, outcomes were modeled independently without competing-risks or joint modeling. Future work will prioritize external validation, non-linear probes, partial fine-tuning, larger genetic cohorts, and competing-risks analyses using more comparable endpoints.

### Future work

We plan to (1) pursue external validation at an additional OMOP/MEDS site; (2) evaluate non-linear probes and partial fine-tuning of CLMBR-T-base on the same tasks; (3) expand the genetic-testing-linked cohort through multi-site collaboration to stabilize the genotype-prediction estimates; and (4) extend the outcomes analysis to a competing-risks framework with endpoint definitions more directly comparable to the existing HCM/DCM risk-stratification literature.

## Data Availability

Data produced in the present study are not publicly available.

## Acknowledgements

We acknowledge the Penn Medicine BioBank (PMBB) for providing data and thank the patient-participants of Penn Medicine who consented to participate in this research program. We would also like to thank the Penn Medicine BioBank team and Regeneron Genetics Center for providing genetic variant data for analysis. The PMBB is approved under IRB protocol# 813913 and supported by Perelman School of Medicine at University of Pennsylvania, a gift from the Smilow family, and the National Center for Advancing Translational Sciences of the National Institutes of Health under CTSA award number UL1TR001878.

